# Telomere length and cardiovascular mortality: a multistate competing risk analysis

**DOI:** 10.1101/2025.07.02.25330776

**Authors:** Hanish P. Kodali, Luisa N. Borrell, Linda Valeri, Heidi E. Jones, Katarzyna E. Wyka

## Abstract

**Background:** Cardiovascular disease (CVD) is the leading cause of mortality in the United States, with substantial economic and health impacts. While traditional risk factors are well studied, non-traditional factors like telomere length (TL), have garnered interest due to mixed findings on their association with CVD-specific mortality. This study investigates the association between TL and CVD-specific mortality, accounting for non-CVD-specific mortality as a competing risk, using a multistate framework.

**Methods:** We conducted a retrospective cohort study using data from the National Health and Nutrition Examination Survey 1999-2002 and 2019 Linked Mortality Files. This study included 6,516 non-institutionalized adults aged 25 years or older. TL was measured using quantitative PCR and analyzed continuously and categorically. We employed a multistate model to evaluate transitions from an event-free state to CVD-specific and non-CVD-specific mortality, estimating cause-specific hazard ratios (HRs) adjusted for sociodemographic and health risk factors.

**Results:** The cumulative incidence function for CVD-specific mortality was significantly higher in the lowest TL quartile than in the highest quartile (Gray test, p < 0.01). Gray’s test was used to compare CIFs across quartiles without applying a Fine and Gray model. We found that a shorter TL was associated with a higher risk of CVD-specific mortality. In the adjusted model, each unit decrease in TL was associated with a 57% higher rate of CVD-specific mortality (HR: 1.57, 95% CI: 1.24-1.98). Adults in the shortest TL quartile had an 88% higher rate of CVD-specific mortality compared with those in the longest TL quartile (HR: 1.88, 95% CI: 1.29-2.72).

**Conclusion:** Our findings suggest a significant inverse association between TL and CVD-specific mortality, highlighting TL as a potential biomarker for CVD risk. The use of a multistate framework provides a comprehensive understanding of competing risks and enhances the robustness of our results. Further studies are needed to validate these findings and explore the underlying mechanisms.

## INTRODUCTION

Cardiovascular disease (CVD) remains the leading cause of mortality in the United States (U.S.).^1^ According to the CDC, approximately 1 in every 5 deaths in the U.S. were attributable to heart disease in 2021.^2^ Additionally, the annual cost of CVD between 2018 and 2019 was estimated at $239.9 billion, encompassing healthcare services, medicines, and lost productivity due to death.^3^ Despite significant advances in understanding the pathophysiology of CVD and targeting traditional risk factors, substantial reductions in mortality have not been achieved. Consequently, there is a growing interest in investigating non-traditional risk factors.^4–6^

One such non-traditional risk factor is telomere length (TL). Several studies have evaluated the role of TL in CVD-specific mortality risk. For instance, a meta-analysis, including 14,491 participants from 18 studies conducted in Asia, Europe, and North America, found that TL was significantly shorter in individuals diagnosed with coronary heart disease and was inversely associated with disease severity.^7^ These findings are corroborated by a Mendelian randomization study conducted in the United Kingdom among a middle-aged population.^8^ Currently, TL is considered a biomarker for future cardiovascular events and a measure of the efficacy of preventive measures against heart disease.^9^ However, findings from studies employing time-to-event analyses are mixed.^10–16^ For example, a study conducted in the U.S. and Germany, involving 12,199 participants with over 12 years of follow-up, found no association between TL and CVD-specific mortality risk.^11^ In contrast, another study conducted in the U.S. on a nationally representative population with 14.4 years of follow-up, did find an inverse association, i.e., a shorter TL was associated with higher mortality rate.^12^

As individuals age, the incidence and prevalence of cardiovascular risk factors and comorbid conditions like diabetes and cancer increase.^17^ These comorbid conditions may share a similar pathophysiology with CVD and may interfere with the occurrence of CVD-specific mortality or modify its probability of onset. These comorbid conditions are labeled as competing events that can hinder the occurrence or modify the probabilities of the event of interest. Studies have shown that ignoring competing risks can produce biased estimates.^18,19^ Therefore, it is crucial to evaluate the association of TL with CVD-specific mortality, while addressing the impact of death due to non-cardiovascular causes as competing risks.

Two methods are often employed for competing risk analysis: cause-specific and Fine-Gray regression models. The first is a cause-specific hazard regression model condition, such as CVD-specific mortality, while censoring all other events, including those who are alive, lost to follow-up, or died due to competing risks (i.e., non-CVD-specific mortality). The second, Fine-Gray regression model, does not censor a participant who experienced a competing event(s) and lets them remain in the risk-set, making it another approach for competing risk analysis.^20^ However, the Fine-Gray model has been shown to have a fundamental mathematical problem, as demonstrated by Austin et al..^21^ Specifically, when using real-life data to estimate the cumulative incidence functions (CIFs) for multiple competing event types, the sum of the estimated CIFs across the event types can exceed 1 (or 100%), thereby violating probability laws.

To accommodate the presence of competing risks, we used a multistate framework that extends traditional competing risk models and provides a flexible framework in which subjects transition between a finite number of states. The multistate model conceptualizes a stochastic process in terms of a set of comprehensive, mutually exclusive states and the transitions among them, accounting for competing events at each transition.^22^ The occurrence of a competing risk is modeled as a transition out of an initial state (e.g., event-free) into a competing risk state (e.g., non-CVD). This transition occurs at the time of the first event.^23^ This approach allows for a more comprehensive understanding of the association between TL and CVD-specific mortality by considering the transitions between different health states, including non-CVD-specific death, as a competing risk. This study aimed to contribute to the understanding of TL as a potential biomarker for CVD mortality in the context of competing risks.

## METHODS

### Data sources

We conducted a retrospective cohort study using two publicly available datasets: National Health and Nutrition Examination Survey (NHANES) 1999-2002^24,25^ and 2019 Linked Mortality Files (LMF).^26^ NHANES, conducted by the National Center for Health Statistics (NCHS), a division of the Centers for Disease Control and Prevention, is a nationally representative survey that collects health-related information annually from 5,000 non-institutionalized civilians across the U.S. The survey includes interviews, physical examinations, and laboratory tests to gather comprehensive data on sociodemographic factors, nutritional information, and health and health related self-reported and objective measurements.^27^

NHANES uses a multistage probability sampling design to generate population-level estimates. This design involves four stages: selecting primary sampling units (usually U.S. counties), sampling census blocks or block combinations, identifying non-institutional group quarters, such as dormitories, and selecting individuals from households or dwelling units. Specific subpopulations, including Mexican-Americans, Black individuals, adolescents (aged 12 to 19 years), and older adults (aged 60 years or older), are oversampled to improve the precision of estimates for these groups.^27^

Participants of the NHANES were linked to the 2019 LMF from the National Death Index (NDI) using a probabilistic matching algorithm.^28^ This algorithm uses information such as social security number, first name, middle initial, last name or surname, father’s surname, birthdate (month, day, and year), sex, state or country of birth, race, and state of residence.^28^

### Outcome

For each participant, the time-to-event was determined from the participant’s date of the interview until either the date of death (for participants who had passed away) or December 31, 2019 (for participants still alive).^26^ Further, among the those passed away, based on the International Classification of Diseases, tenth revision (ICD-10), cause-specific of death was determined using the underlying cause of death (UCOD).^29^ When UCOD was coded I00 to I09, I11, I13, I20 to I51, and I60 to I69 as per ICD-10, we labelled the event as CVD-specific mortality, while other deaths were labelled as non-CVD-specific mortality.

### Exposures

Among participants aged 20 years or older who provided DNA and consented to future genetic research, TL measurements were obtained from blood samples in the 1999-2002 NHANES. Using the quantitative polymerase chain reaction (PCR) method, TL was assessed and compared with a standard reference DNA using the T/S ratio.^24,25^ Consistent with previous studies, the T/S ratio was converted to kilobase pairs (kbp) using the following formula: ((3,274 + 2,413 * (T/S))/1,000). TL measurements ranged from 4.2 to 26 kbp, with a mean of 5.71 kbp and a standard deviation of 0.67. In this analysis, TL was treated as a continuous and categorical variable. To reflect a positive association with mortality risk, we multiplied TL by -1. Additionally, to quantify the effect magnitude, TL was categorized into quartiles (Q1, Q2, Q3, and Q4) with cutoff points of 5.27, 5.63, and 6.04 kbp.

### Covariates

Based on a literature review^16,30–36^ and considering potential covariates, we considered baseline sociodemographic characteristics and health risks in the analyses. Age was provided as a continuous variable (in years) and further categorized into three groups: young (25-44 years), middle (45-64 years), and older adults (65 years or older). Self-reported sex was considered as recorded, female or male. We used self-reported race/ethnicity data, as provided by NHANES, and focused only on non-Hispanic White, non-Hispanic Black, (hereafter referred to as White and Black, respectively), and Mexican American. Self-reported education was coded as follows: less than high school (less than 9^th^ grade, 9-11^th^ grade, and 12^th^ grade with no diploma), high school (high school graduate/GED or equivalent), some college (or AA degree), and college graduate or above. Smoking status was determined based on two NHANES survey questions: “Have you smoked at least 100 cigarettes in your entire life?” and “Do you now smoke cigarettes?” Participants will be categorized as ‘never smoked’ (responded “No” to the first question), ‘former smoker’ (responded “Yes” to the first and “No” to the second question), or ‘current smoker’ (responded “Yes” to both questions).

### Study population

Of the 21,004 U.S. residents who participated in the 1999–2002 NHANES, we excluded participants ineligible for mortality follow-up due to insufficient identifying data or age under 18 (n = 9,572), those younger than 20 or who declined DNA consent for genetic research (n = 3,610), individuals under 25 (n = 758), participants with race/ethnicity listed as “Other,” “Other Hispanics,” or missing (n = 529), and those missing data for education (n = 9) or smoking status (n = 6). After these exclusions, the analytical sample size was 6,520 participants. Among these participants, 2,164 had died as of December 31, 2019 (Supplementary Figure 1). The average follow-up duration of the cohort was 16.73 years (SE= 0.08), ranging from 0.08 to 20.83 years.

### Statistical analysis

We reported means and standard errors (SE) for continuous variables, including TL, and frequencies and proportions for categorical variables by event type. For competing risks, we estimated the cumulative incidence function (CIF) for both CVD- and non-CVD-specific mortality events. The CIF estimates the probability of experiencing a specific event type over time, while accounting for the presence of competing risks. Further, CIF was calculated separately for each TL quartile to observe the probability of CVD-specific mortality, treating non-CVD mortality as a competing risk. To compare the cumulative incidence of CVD-specific events across different TL quartiles, Gray’s test, a non-parametric test for equality of CIFs, was used. This comparison was conducted independently of a multistate or Fine and Gray model.^37,38^

In our multistate model, we defined event-free state, CVD death, and non-CVD-specific death. A change in the state is called a transition. All participants were event-free, with some transitioning to CVD-death (transition 1) and others to non-CVD-death (transition 2). Both death states absorbing (terminal) states because no further transitions can occur from these states. Cause-specific hazards determine the transition intensity from non-event to CVD- or non-CVD-specific mortality. Each transition rate was represented by a separate model equivalent to a Cox model that focused on the endpoint of the transition. We fitted two models: an unadjusted and an adjusted for sociodemographic characteristics and health risks. Proportional hazard assumptions were assessed using Schoenfeld residuals.^39^ We reported hazard ratios (HRs) and 95% confidence intervals (CIs), which were interpreted similarly to those in a Cox model.

Furthermore, we conducted a sensitivity analysis comparing estimates across three methods: the Cox proportional hazards model, the multistate competing risk model, and the Fine and Gray sub-distribution hazards model. Because the Fine and Gray method does not accommodate survey weights, this analysis used unweighted data. While Fine and Gray’s estimates are not directly comparable to those from the other two methods, this comparison allows us to examine the consistency in the magnitude and direction of the estimates.

Data management and analysis were conducted in R using the “tidyverse”^40^ package for data manipulation and “srvyr”^41^ to account for the complex sampling design of NHANES. The Aalen-Johansen estimator for CIF calculation was employed using the “cmprsk”^42^ package, and the “mstate”^43^ package was used for multistate modeling. Statistical significance was set at p < 0.05.

## RESULTS

Table 1 presents the baseline characteristics by event type i.e. event-free, CVD-specific, and non-CVD-specific mortality. The total sample was predominantly younger (45.1% aged 25-44 years), White (82.8%), and never smokers (48.6%). Older adults (65 years or older) were more likely to be in CVD-specific (64.8%) and non-CVD-specific (55.6%) mortality groups than in the event-free group (19.3%) (p < 0.001). Lower education levels were more prevalent in the CVD-specific mortality group (37.1%) than in the event-free group (20.1%) (p < 0.001). Current smokers were more common in the non-CVD-specific mortality group (28.3%) than in the CVD-specific mortality (18.9%) and event-free (23.6%) groups (p < 0.001). Distribution by sex was similar across the groups. Mean telomere length (TL) was shorter in the CVD-specific mortality group (5.46 kbp) than in the event-free group (5.77 kbp) (p < 0.001). The lowest TL quartile (Q1) had the highest CVD-specific mortality (40.7%) compared with the highest quartile (Q4) (12.3%) (p < 0.001).

**Table 1:** Total and event type distribution for selected characteristics among adults 25 years or older who participated in National Health and Nutrition Examination Survey, 1999-2002

| Characteristic | Total<br>(n = 6,520) | Event Free<br>(n = 4,356) | CVD-Specific<br>Death<br>(n = 696) | Non-CVD-Specific Death<br>(n = 1,468) | p-value<sup>1</sup> |
| --- | --- | --- | --- | --- | --- |
| Age Groups, % (SE) |  |  |  |  | <0.001 |
| Young (25-44 years) | 45.05 (0.01) | 56.72 (0.01) | 8.15 (0.02) | 10.92 (0.01) |  |
| Adult (45-64 years) | 35.66 (0.01) | 37.05 (0.01) | 27.06 (0.03) | 33.47 (0.01) |  |
| Older (65 years or older) | 19.30 (0.01) | 6.23 (0.00) | 64.79 (0.03) | 55.61 (0.02) |  |
| Sex, % (SE) |  |  |  |  | 0.043 |
| Male | 48.71 (0.01) | 47.93 (0.01) | 53.78 (0.02) | 49.82 (0.01) |  |
| Female | 51.29 (0.01) | 52.07 (0.01) | 46.22 (0.02) | 50.18 (0.01) |  |
| Race/Ethnicity, % (SE) |  |  |  |  | <0.001 |
| Non-Hispanic White | 82.77 (0.01) | 81.63 (0.01) | 86.31 (0.02) | 86.15 (0.01) |  |
| Non-Hispanic Black | 10.21 (0.01) | 10.20 (0.01) | 10.95 (0.02) | 9.93 (0.01) |  |
| Mexican American | 7.02 (0.01) | 8.17 (0.01) | 2.74 (0.01) | 3.92 (0.01) |  |
| Education, % (SE) |  |  |  |  | <0.001 |
| Less than High School | 20.08 (0.01) | 15.72 (0.01) | 37.13 (0.03) | 31.33 (0.02) |  |
| High School | 26.34 (0.01) | 25.60 (0.01) | 25.47 (0.03) | 29.93 (0.02) |  |
| Some College | 27.38 (0.01) | 28.32 (0.01) | 21.81 (0.02) | 25.82 (0.02) |  |
| College | 26.20 (0.02) | 30.35 (0.02) | 15.59 (0.02) | 12.92 (0.02) |  |
| Smoking Status, % (SE) |  |  |  |  | <0.001 |
| Never | 48.61 (0.01) | 51.97 (0.01) | 42.32 (0.02) | 36.87 (0.01) |  |
| Former | 27.76 (0.01) | 25.00 (0.01) | 38.81 (0.02) | 34.81 (0.02) |  |
| Current | 23.63 (0.01) | 23.03 (0.01) | 18.88 (0.02) | 28.32 (0.02) |  |
| Telomere Length (kbs),<br>Mean (SE) | 5.77 (0.04) | 5.86 (0.04) | 5.46 (0.04) | 5.52 (0.04) | <0.001 |
| Telomere Length (in<br>quartile, % (SE) |  |  |  |  | <0.001 |
| Q1: < 5.27 | 21.02 (0.02) | 15.44 (0.02) | 40.73 (0.04) | 36.39 (0.03) |  |
| Q2: 5.27 - 5.63 | 24.69 (0.01) | 23.90 (0.01) | 26.24 (0.02) | 27.43 (0.02) |  |
| Q3: 5.63 - 6.04 | 26.17 (0.01) | 27.88 (0.01) | 20.74 (0.02) | 21.18 (0.01) |  |
| Q4: > 6.04 | 28.12 (0.02) | 32.78 (0.02) | 12.29 (0.03) | 15.01 (0.02) |  |
| <sup>1</sup> chi-squared test with Rao & Scott's second-order correction; Wilcoxon rank-sum test for complex survey samples |  |  |  |  |  |
| NOTE: CVD-Cardiovascular |  |  |  |  |  |

Figure 2 presents the CIF estimates for both CVD- and non-CVD-specific mortality at 50-, 100-, 150-, 200- and 250-month follow-up periods. The 250-month probability of CVD-specific mortality was 4.9% (95% CI: 3.9%, 6.1%, Table 2), and for non-CVD-specific mortality was 14.4% (95% CI: 12.1%, 17.2%, Table 2). These results suggest that the risk of CVD-specific mortality is lower than that of mortality from other causes within this timeframe.

**Figure 1:**
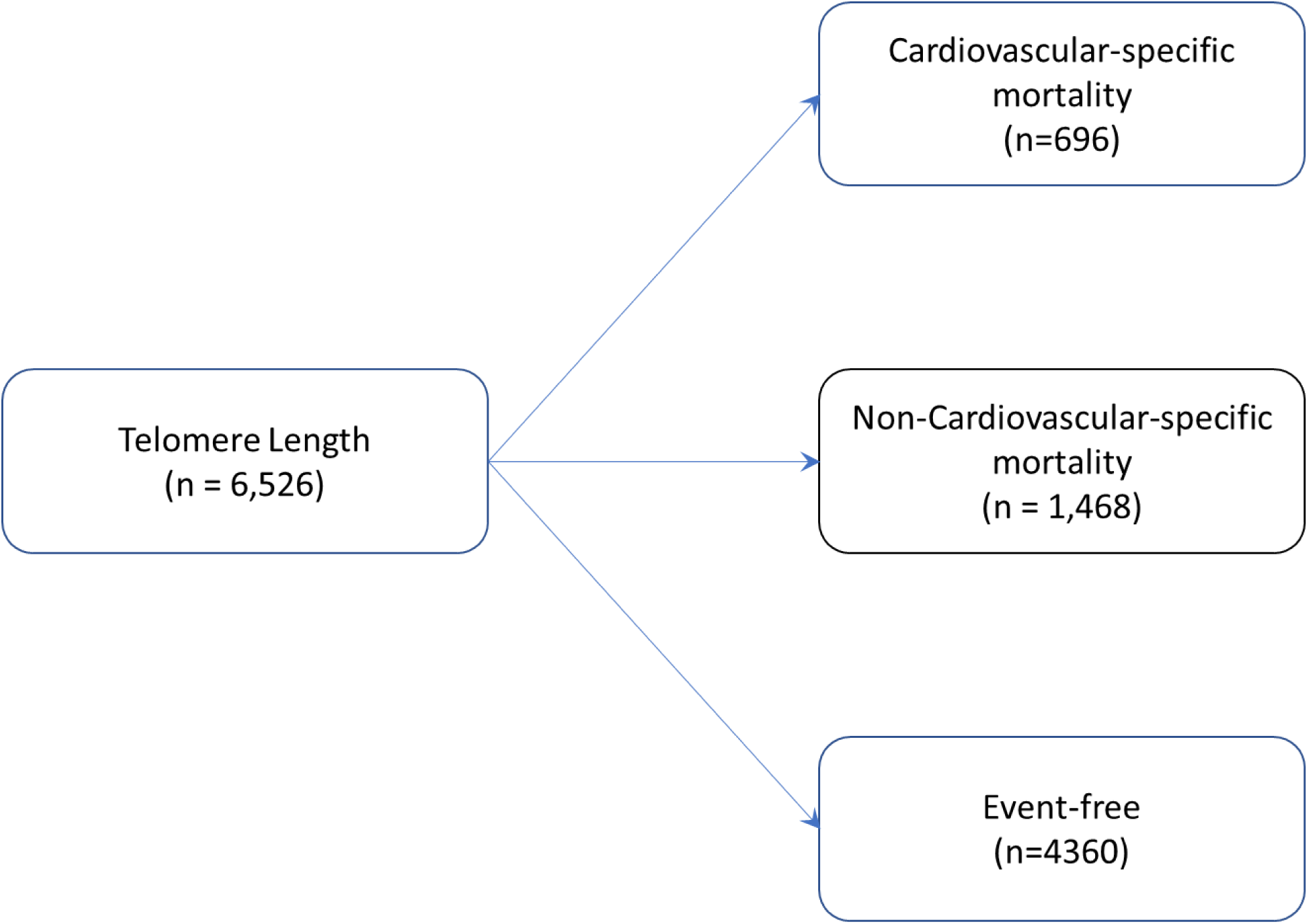
A schematic representation of the multistate model showing the different states and the possible transitions between these states (boxes representing states and arrows indicating possible transitions): National Health and Nutrition Examination Survey, 1999-2002, and 2019 Linked Mortality File

**Figure 2:**
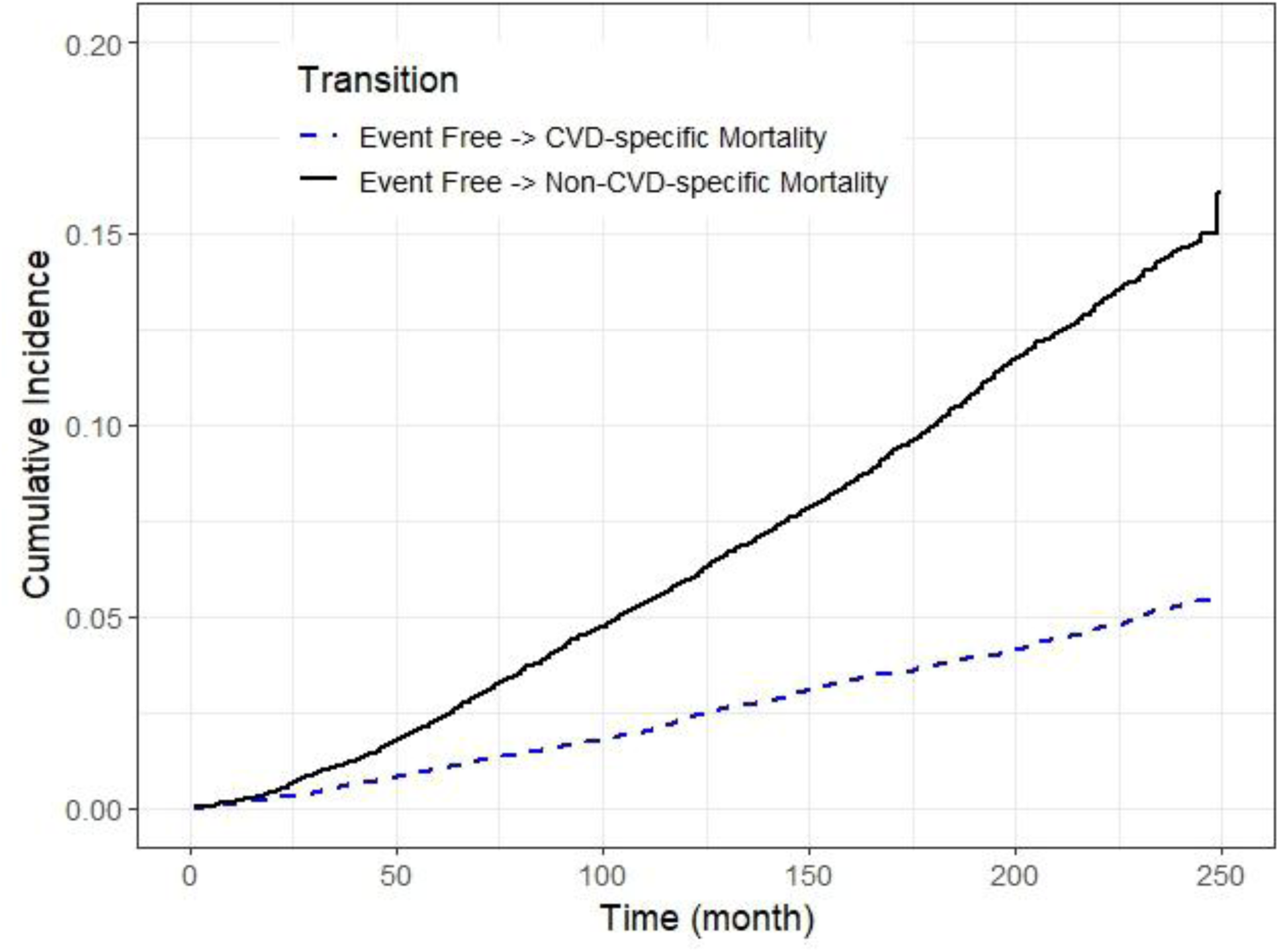
Cumulative incidence functions for cardiovascular (CVD)-, and non-CVD-specific mortality over time: National Health and Nutrition Examination Survey, 1999-2002, and 2019 Linked Mortality File

**Table 2:** Cumulative incidence function estimates (CIF) % and 95% confidence intervals (CI) for cardiovascular (CVD)-, and non-CVD-specific mortality among adults 25 years or older: National Health and Nutrition Examination Survey, 1999-2002, and 2019 Linked Mortality File

|  | **Month 50** | **Month 100** | **Month 150** | **Month 200** | **Month 250** |
| --- | --- | --- | --- | --- | --- |
| **Event Type** | **CIF% (95% CI)** | **CIF% (95% CI)** | **CIF% (95% CI)** | **CIF% (95% CI)** | **CIF% (95% CI)** |
| CVD-Specific Death | 0.8 (0.6, 1.1) | 1.7 (1.3, 2.2) | 2.9 (2.3, 3.7) | 3.8 (3.1, 4.8) | 4.9 (3.9, 6.1) |
| Non-CVD-Specific Death | 1.8 (1.5, 2.1) | 4.6 (4.0, 5.3) | 7.5 (6.5, 8.6) | 10.8 (9.5, 12.4) | 14.4 (12.1, 17.2) |

Figure 3 illustrates the CIF curves for CVD-specific and non-CVD-specific mortality by TL quartiles. Accounting for censoring and competing events, the 250-month cumulative incidence of CVD-specific mortality was 19% (95% CI: 17%, 21%, Table 3) in the lowest TL quartile (Q1) compared with 5% (95% CI: 3.9%, 6.3%, Table 3) in the highest TL quartile (Q4). Gray’s test indicated a statistically significant difference in CIFs among these quartiles (p < 0.01), suggesting that individuals in the lowest TL quartile had a higher probability of CVD-specific mortality over time than those in the highest TL quartile (Table 3).

**Figure 3:**
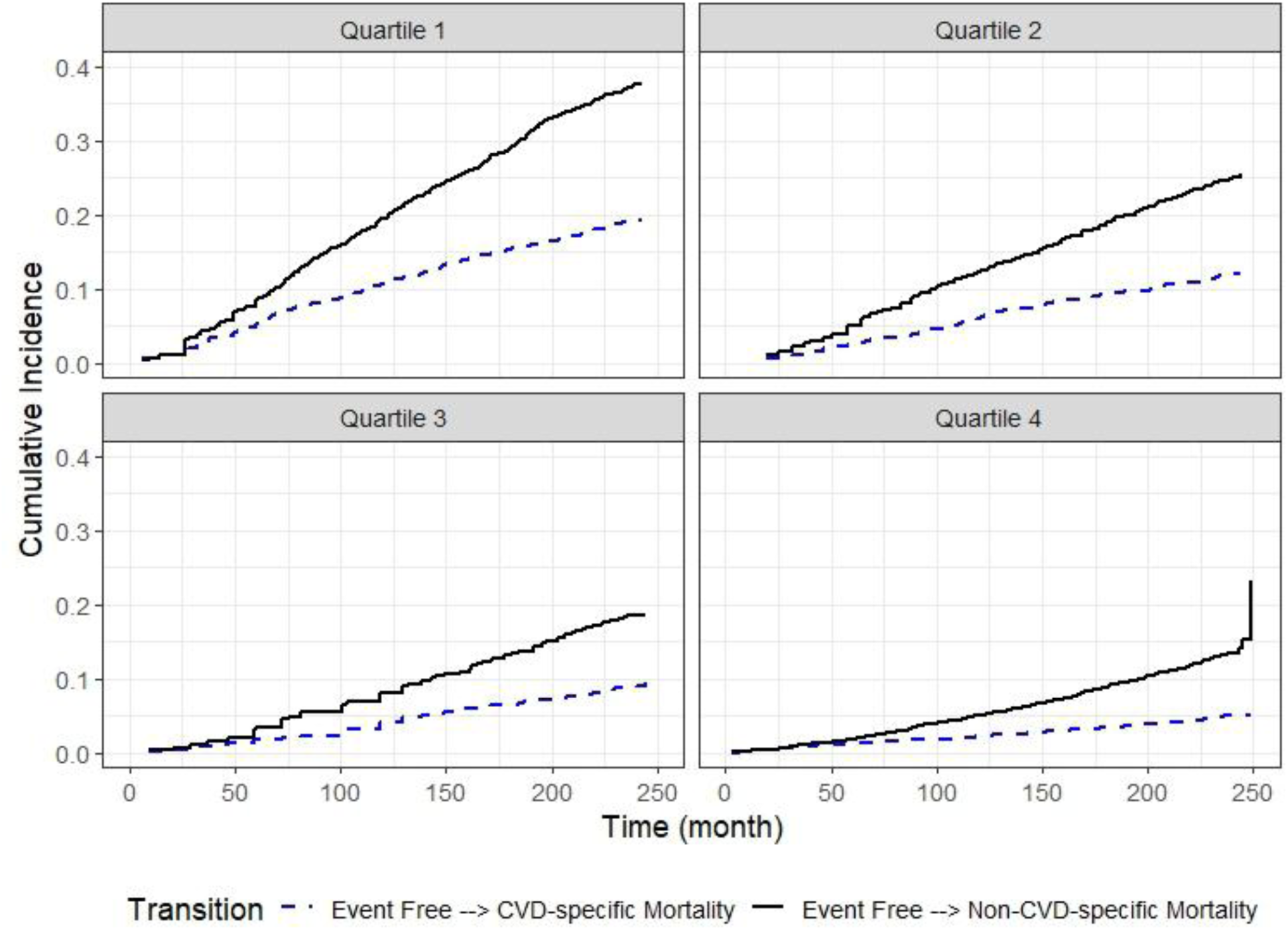
Cumulative incidence functions for cardiovascular (CVD)-, and non-CVD-specific mortality over time, stratified by TL quartiles: National Health and Nutrition Examination Survey, 1999-2002, and 2019 Linked Mortality File

**Table 3:** Cumulative incidence function estimates (CIF) % and 95% confidence intervals (CI) for cardiovascular (CVD)-, and non-CVD-specific mortality among adults 25 years or older stratified by quartiles of the telomere length: National Health and Nutrition Examination Survey, 1999-2002, and 2019 Linked Mortality File

|  | Month 50 | Month 100 | Month 150 | Month 200 | Month 250 | p-value<sup>1</sup> |
| --- | --- | --- | --- | --- | --- | --- |
| Characteristic | CIF% (95% CI) | CIF% (95% CI) | CIF% (95% CI) | CIF% (95% CI) | CIF% (95% CI) |  |
| **Cardiovascular (CVD)-Specific Death** |  |  |  |  |  |  |
| Telomere Length (in quartile) |  |  |  |  |  | <0.001 |
| Q1: < 5.27 | 4.4 (3.4, 5.4) | 8.7 (7.4, 10.0) | 13.0 (12.0, 15.0) | 16.0 (15.0, 18.0) | 19.0 (17.0, 21.0) |  |
| Q2: 5.27 - 5.63 | 2.1 (1.5, 2.9) | 4.7 (3.7, 5.8) | 7.9 (6.7, 9.3) | 9.8 (8.4, 11.0) | 12.0 (10.0, 14.0) |  |
| Q3: 5.63 - 6.04 | 1.4 (1.0, 2.1) | 2.9 (2.2, 3.9) | 5.5 (4.5, 6.7) | 7.3 (6.1, 8.6) | 9.5 (7.8, 11.0) |  |
| Q4: > 6.04 | 0.9 (0.6, 1.6) | 1.8 (1.2, 2.5) | 2.7 (2.0, 3.6) | 3.9 (3.0, 4.9) | 5.0 (3.9, 6.2) |  |
| **Non-CVD-Specific Death** |  |  |  |  |  |  |
| Telomere Length (in quartile) |  |  |  |  |  | <0.001 |
| Q1: < 5.27 | 7.0 (5.8, 8.3) | 16.0 (14.0, 18.0) | 24.0 (22.0, 27.0) | 33.0 (31.0, 35.0) | 38.0 (35.0, 40.0) |  |
| Q2: 5.27 - 5.63 | 4.0 (3.1, 5.0) | 10.0 (8.9, 12.0) | 16.0 (14.0, 17.0) | 21.0 (19.0, 23.0) | 25.0 (23.0, 28.0) |  |
| Q3: 5.63 - 6.04 | 2.4 (1.7, 3.2) | 6.4 (5.3, 7.6) | 11.0 (9.1, 12.0) | 15.0 (14.0, 17.0) | 19.0 (17.0, 21.0) |  |
| Q4: > 6.04 | 1.5 (1.0, 2.2) | 4.0 (3.1, 5.0) | 6.6 (5.5, 7.9) | 10.0 (8.8, 12.0) | 23.0 (9.7, 40.0) |  |
| <sup>1</sup> Gray's Test |  |  |  |  |  |  |

### Cardiovascular-specific mortality

In the adjusted model, for the transition from an event-free state to CVD-specific mortality, with every unit decrease in TL, the rate of death due to CVD-specific mortality was 1.57 times higher (95% CI: 1.24, 1.98, Table 4, Figure 4). When considering TL quartiles, participants in the shortest TL quartile had a 1.88 times higher rate of dying (95% CI: 1.29, 2.72) than those in the longest TL quartile (Table 4, Figure 4).

**Figure 4:**
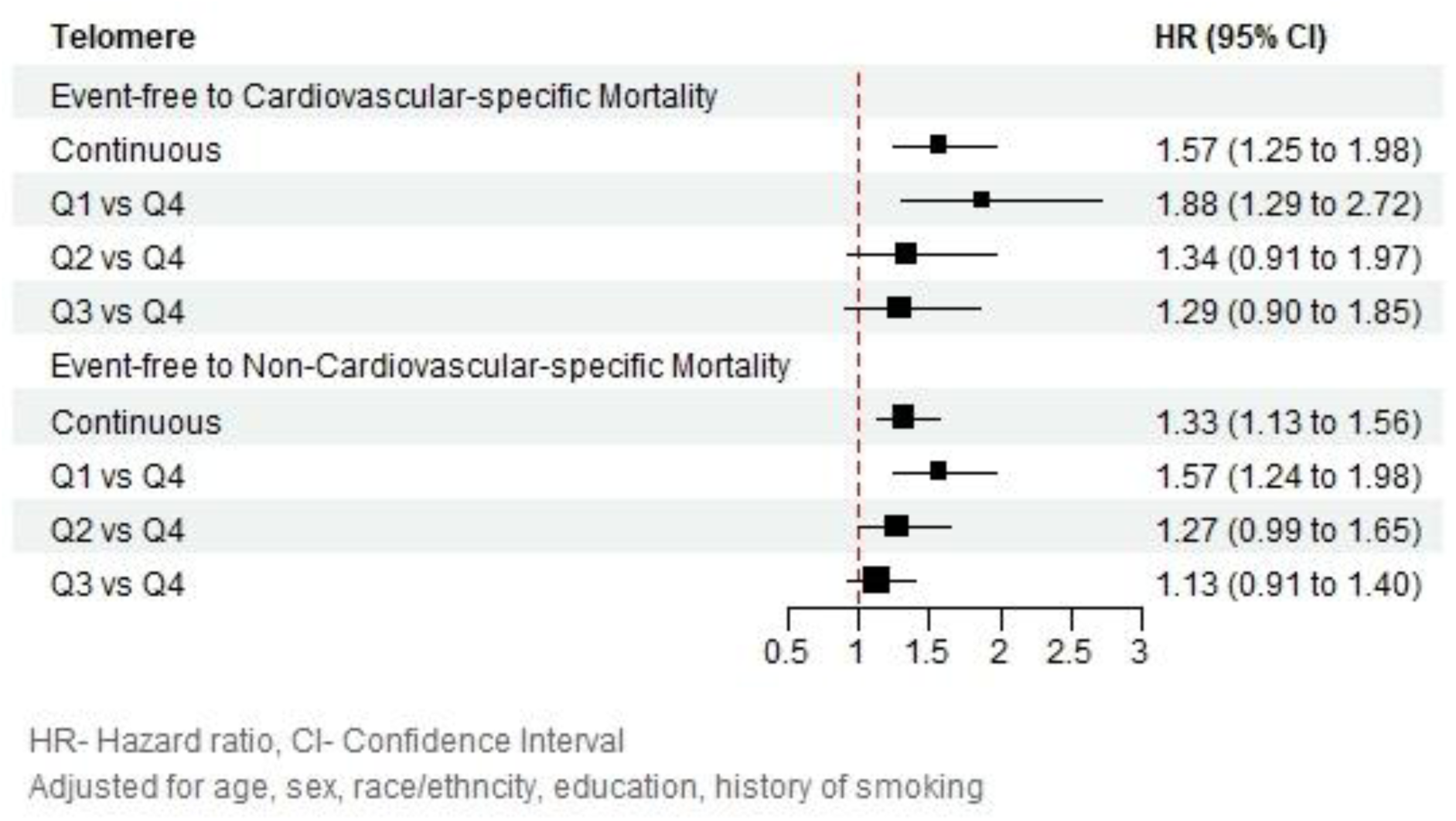
Adjusted hazards ratios (HR) and 95% confidence intervals (CI) for leucocyte telomere length on cardiovascular-specific Mortality and non-cardiovascular-specific Mortality: National Health and Nutrition Examination Survey, 1999-2002, and 2019 Linked Mortality File

**Table 4:** Hazard ratios (HR) and 95% confidence intervals (CI) for transitions between states for leucocyte telomere length on cardiovascular (CVD)-, and non-CVD-specific mortality risks: National Health and Nutrition Examination Survey, 1999-2002, and 2019 Linked Mortality File

|  |  | **Model 1<sup>2</sup>** | **Model 2<sup>3</sup>** |
| --- | --- | --- | --- |
| **Transition** | **Variable** | **HR (95% CI)** | **HR (95% CI)** |
| Event-free to CVD-specific mortality | Telomere (kb)<sup>1</sup> | 3.45 (2.56, 4.65) | 1.57 (1.24, 1.98) |
|  | Quartiles |  |  |
|  | Q1: < 5.27 | 5.45 (3.48, 8.55) | 1.88 (1.29, 2.72) |
|  | Q2: 5.27 - 5.63 | 2.66 (1.74, 4.08) | 1.34 (0.91, 1.97) |
|  | Q3: 5.63 - 6.04 | 1.88 (1.25, 2.82) | 1.29 (0.90, 1.85) |
|  | Q4: > 6.04 | 1.00 | 1.00 |
| Event-free to non-CVD-specific mortality | Telomere (kb)<sup>1</sup> | 2.60 (2.06, 3.28) | 1.33 (1.13, 1.56) |
|  | Quartiles |  |  |
|  | Q1: < 5.27 | 4.02 (2.96, 5.46) | 1.57 (1.24, 1.98) |
|  | Q2: 5.27 - 5.63 | 2.28 (1.73, 3.02) | 1.27 (0.99, 1.65) |
|  | Q3: 5.63 - 6.04 | 1.57 (1.23, 2.01) | 1.13 (0.91, 1.40) |
|  | Q4: > 6.04 | 1.00 | 1.00 |
| <sup>1</sup>The continuous measure of telomere length was multiplied by -1 to reflect increased risk of mortality associated with shorter telomere length |  |  |  |
| <sup>2</sup>Model 1- Unadjusted models |  |  |  |
| <sup>3</sup>Model 2- Adjusted for age, sex, race/ethnicity, education, smoking status |  |  |  |

### Non-Cardiovascular-specific mortality

In the adjusted model, for the transition from the event-free state to non-CVD-specific mortality, with every unit decrease in TL, the rate of death due to non-CVD-specific mortality was 1.33 times higher (95% CI: 1.13, 1.56, Table 4, Figure 4). Further, for TL quartiles, participants in the shortest TL quartile had a 1.57 times higher rate of death (95% CI: 1.24, 1.98) from all-causes other than CVD than those in the longest TL quartile (Table 4, Figure 4).

For sensitivity analysis, results from the Cox proportional hazards model, the multistate competing risk model, and the Fine and Gray sub-distribution hazards model showed consistent direction and magnitude of associations between telomere length and both CVD-specific and non-CVD-specific mortality. Full results are provided in the Appendix (Supplement Table 1).

## DISCUSSION

Among U.S. adults aged 25 years or older, we found an inverse association between TL and CVD-specific mortality risk after accounting for competing mortality events. Notably, the association was particularly strong among individuals with the lowest quartile compared with those with the highest quartile.

While we cannot make direct comparisons, our findings align with those of previous studies,^12–14,30,44^ indicating that a short TL is associated with higher CVD-specific mortality. However, a recent Mendelian randomization (MR) study conducted using the UK Biobank data, involving 472,174 individuals aged 40 to 69 years, reported a significant association between longer TL and various cardiovascular disease conditions but did not find a significant association with cardiovascular death.^8^ We hypothesize two main reasons for this difference in results. First, the study populations in the MR study were predominantly European White individuals, which may have introduced some population-specific genetic variations that could influence the association between TL and cardiac death. Current research suggests that genetic determinants of TL can vary across populations, potentially due to differences in allele frequencies and environmental interactions.^45^ This lack of diversity in study populations could result in differences in how genetic factors related to TL impact cardiovascular health outcomes.^46,47^ Therefore, findings from a predominantly European White population may not fully capture the genetic and environmental dynamics that influence TL and cardiac risk in more diverse populations.^46,47^ Additionally, environmental and lifestyle factors that differ between populations could interact with genetic factors to influence TL and cardiovascular outcomes. In contrast, our study involved a nationally representative sample of the U.S. adult population, which is known for its racial and ethnic diversity. This diversity might have contributed to the observed associations. This is consistent with the findings of Acosta et al.,^48^ who utilized cause-of-death data from the World Health Organization mortality database (2000–2016) and compared CVD mortality trends in the U.S. to those in 17 other high life expectancy countries (HLCs). Their trend analysis revealed that the U.S. has a less favorable trajectory of CVD-specific mortality relative to other HLCs, including the United Kingdom. This trend is influenced by factors such as obesity, alcohol-related mortality, and growing socioeconomic inequalities in the U.S. Second, the MR study excluded adults aged 69 years or older, which means that the study did not capture many CVD deaths in individuals older than 69 years. This exclusion might have affected the results, as CVD deaths are more common in older age groups. In contrast, our study included adults aged 25 years or older, with a maximum follow-up of 20 years, which might have allowed us to capture a broader range of cardiac deaths. To the best of our knowledge, no previous study has used a multistate framework to evaluate this association, making our approach novel.

The analysis of TL in relation to CVD-specific mortality, considering non-CVD mortality as a competing risk, can be significantly improved by employing a multistate framework. Traditional methods like the cause-specific hazard model and the Fine-Gray sub-distribution hazard model, have notable limitations. The cause-specific hazard model, which censors competing events, can lead to biased estimates if the competing risk shares common risk factors with the primary event.^49^ The Fine-Gray model, while addressing this issue, can produce probabilities exceeding 1, lacks straightforward integration with complex survey designs, and does not easily interpret absolute risks.^21^ However, multistate models provide a robust alternative by conceptualizing competing risks within a unified framework, allowing simultaneous modeling of multiple event transitions.^50^ This approach not only yields interpretable absolute risk measures, such as the probability of being in a specific state over time and the mean time spent in each state, but also accounts for the dependencies among competing risks. By directly modeling transition intensities (which correspond to cause-specific hazards), multistate models avoid the need for stringent assumptions and provide a comprehensive view of the disease process, including intermediate events. Moreover, in contrast to the Fine-Gray method, multistate models offer a more general framework that accommodates a variety of competing and intermediate events, thereby enhancing the flexibility and depth of the analysis. The multistate framework’s ability to provide a coherent, probabilistic interpretation of transition probabilities makes it superior for complex time-to-event data analysis involving competing risks.^23^ Thus, using multistate models leads to more accurate and interpretable estimates, offering deeper insights into the relationship between TL and CVD-specific mortality.

### Limitations

Future studies should address several limitations observed in this study. First, a notable proportion of participants did not provide DNA samples or consent for future genetic research, leading to a significant amount of missing TL data. In fact, upon comparing the characteristics of the participants included in the analysis to those excluded due to missing TL data, we observed that the excluded group was predominantly female and more likely to be of Black ethnicity (Supplement Table 2). Second, measurement errors could also occur in the assessment of covariates such as sociodemographic variables and health risks. Inaccurate measurement of these covariates can lead to residual confounding factors. Third, the collection of TL and covariate information at baseline restricted our ability to consider the impact of time-varying changes in these variables. Lastly, the possibility of misclassification of the cause of death (CVD-specific vs. non-CVD-specific) cannot be disregarded and it could further introduce bias, over- or underestimating the number of CVD-specific deaths, and affecting the CIF and cause-specific hazard ratios.^51^ This misclassification can obscure the true relationship between TL and CVD-specific mortality.

### Strengths

Despite these limitations, our study has several strengths. First, the use of NHANES, a nationally representative population sample, allows for the generalizability of our findings to the U.S. population aged 25 years or older. However, as telomere length data were only available for a subset of participants, our results are generalizable to this subset under the assumption that it reflects the broader population, after accounting for the survey’s complex design. The comprehensive nature of the dataset, including detailed information on TL, sociodemographic variables, and risk factors, further supports the validity of our analysis. Second, this study represents the first application of a multi-state analysis using a Markov model to estimate the association between TL and CVD-specific mortality while accounting for non-CVD-specific mortality as a competing event within a nationally representative sample. This approach enables a nuanced analysis of competing risks, offers interpretable absolute risk measures, and provides a comprehensive view of disease progression by modeling multiple event transitions. The multistate model enhances our understanding of the relationship between TL and CVD-specific mortality more thoroughly than previous methods. Additionally, the inclusion of mortality status in a substantial proportion of participants, with up to 20 years of follow-up, strengthens the robustness of our findings. The extended follow-up period ensured that a high number of events were recorded.

## CONCLUSIONS

In this study, we demonstrated a significant inverse association between TL and CVD-specific mortality in a nationally representative sample of U.S. adults. By employing a multistate framework that accounts for competing risks, we provide a nuanced analysis of the relationship between TL and CVD-specific mortality, highlighting the increased risk associated with shorter TL. These findings suggest that TL could serve as a valuable biomarker for predicting CVD-specific mortality, offering potential insights for future preventive strategies and risk assessments. Our study contributes to the growing body of evidence on non-traditional risk factors for CVD and underscores the importance of considering competing risks in epidemiological analyses.

## Data Availability

This study used publicly available de-identified data- National Health and Nutrition Examination Survey (NHANES), and 2019 Public-use Linked Mortality Files

## Supplement Material

**Supplement Figure 1:**
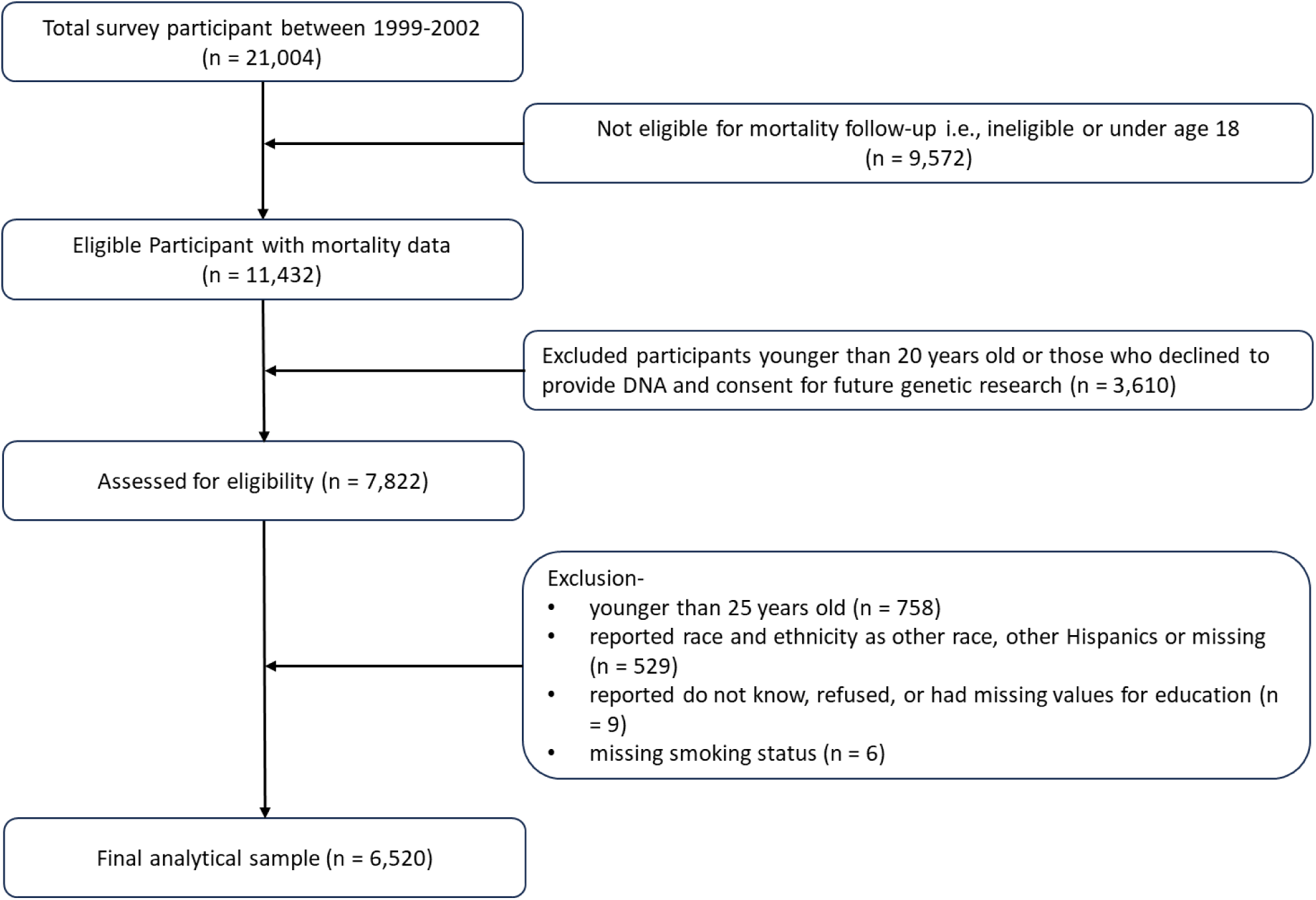
Flowchart depicting sample selection: National Health and Nutrition Examination Survey, 1999-2002

**Supplement Table 1:** Cardiovascular (CVD) - Specific Mortality Risks Across Cox, Multistate, and Fine and Gray Models: National Health and Nutrition Examination Survey, 1999-2002, and 2019 Linked Mortality File

|  | **Cause-Specific**<br>**Cox**<br>**Proportional**<br>**Hazards Model** | **Multistate**<br>**Competing Risk**<br>**Model** | **Fine and Gray**<br>**Subdistribution**<br>**Model** |
| --- | --- | --- | --- |
|  | **HR (95% CI)** | **HR (95% CI)** | **SHR (95% CI)** |
| Telomere (kb)<sup>1</sup> | 1.53 (1.31, 1.79) | 1.53 (1.31, 1.78) | 1.53 (1.31, 1.79) |
| Quartiles |  |  |  |
| Q1: < 5.27 | 1.91 (1.47, 2.49) | 1.91 (1.46, 2.48) | 1.91 (1.46, 2.49) |
| Q2: 5.27 - 5.63 | 1.43 (1.09, 1.88) | 1.43 (1.08, 1.88) | 1.43 (1.08, 1.89) |
| Q3: 5.63 - 6.04 | 1.41 (1.06, 1.87) | 1.39 (1.05, 1.85) | 1.40 (1.05, 1.88) |
| Q4: > 6.04 | 1.00 | 1.00 | 1.00 |
| <sup>1</sup>The continuous measure of telomere length was multiplied by –1 to reflect increased risk of mortality associated with shorter telomere length |  |  |  |
| Models adjusted for age, sex, race/ethnicity, education, smoking status |  |  |  |
| Cause-Specific Cox Proportional Hazards Model: Displays hazard ratios (HRs) and 95% confidence intervals (CIs) estimated by the Cox model |  |  |  |
| Multistate Competing Risk Model: Presents hazard ratios (HRs) and 95% CIs from the multistate competing risk approach |  |  |  |
| Fine and Gray Subdistribution Model: Shows subdistribution hazard ratios (SHRs) and 95% CIs from the Fine and Gray model, unweighted for survey design |  |  |  |

**Supplement Table 2:**
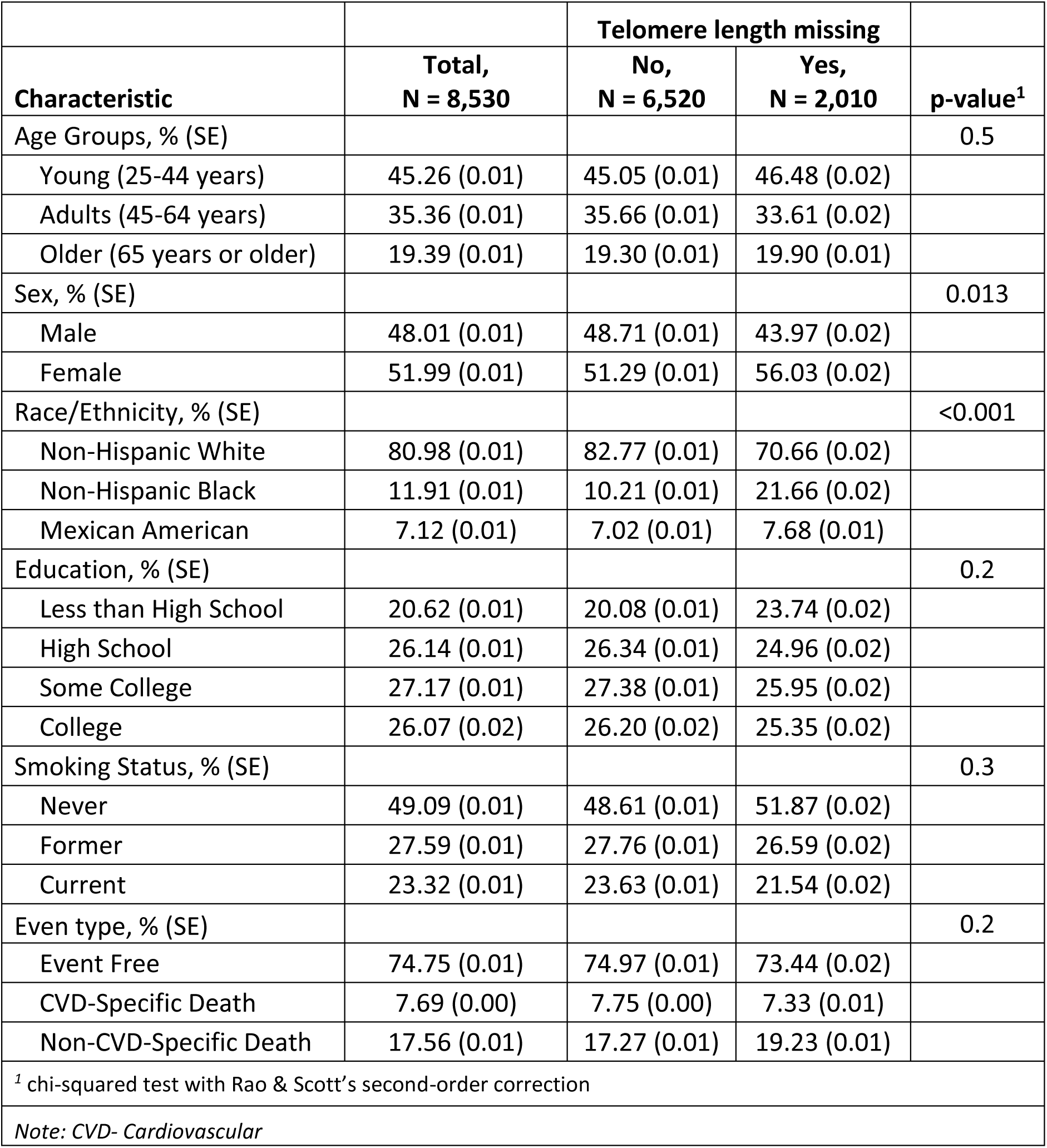
Comparison of selected characteristics among adults 25 years or older participants included and excluded due to missing telomere length information: National Health and Nutrition Examination Survey, 1999-2002, and 2019 Linked Mortality File

|  |  | Telomere length missing |  |  |
| --- | --- | --- | --- | --- |
| Characteristic | Total,<br>N = 8,530 | No,<br>N = 6,520 | Yes,<br>N = 2,010 | p-value<sup>1</sup> |
| Age Groups, % (SE) |  |  |  | 0.5 |
| Young (25-44 years) | 45.26 (0.01) | 45.05 (0.01) | 46.48 (0.02) |  |
| Adults (45-64 years) | 35.36 (0.01) | 35.66 (0.01) | 33.61 (0.02) |  |
| Older (65 years or older) | 19.39 (0.01) | 19.30 (0.01) | 19.90 (0.01) |  |
| Sex, % (SE) |  |  |  | 0.013 |
| Male | 48.01 (0.01) | 48.71 (0.01) | 43.97 (0.02) |  |
| Female | 51.99 (0.01) | 51.29 (0.01) | 56.03 (0.02) |  |
| Race/Ethnicity, % (SE) |  |  |  | <0.001 |
| Non-Hispanic White | 80.98 (0.01) | 82.77 (0.01) | 70.66 (0.02) |  |
| Non-Hispanic Black | 11.91 (0.01) | 10.21 (0.01) | 21.66 (0.02) |  |
| Mexican American | 7.12 (0.01) | 7.02 (0.01) | 7.68 (0.01) |  |
| Education, % (SE) |  |  |  | 0.2 |
| Less than High School | 20.62 (0.01) | 20.08 (0.01) | 23.74 (0.02) |  |
| High School | 26.14 (0.01) | 26.34 (0.01) | 24.96 (0.02) |  |
| Some College | 27.17 (0.01) | 27.38 (0.01) | 25.95 (0.02) |  |
| College | 26.07 (0.02) | 26.20 (0.02) | 25.35 (0.02) |  |
| Smoking Status, % (SE) |  |  |  | 0.3 |
| Never | 49.09 (0.01) | 48.61 (0.01) | 51.87 (0.02) |  |
| Former | 27.59 (0.01) | 27.76 (0.01) | 26.59 (0.02) |  |
| Current | 23.32 (0.01) | 23.63 (0.01) | 21.54 (0.02) |  |
| Event type, % (SE) |  |  |  | 0.2 |
| Event Free | 74.75 (0.01) | 74.97 (0.01) | 73.44 (0.02) |  |
| CVD-Specific Death | 7.69 (0.00) | 7.75 (0.00) | 7.33 (0.01) |  |
| Non-CVD-Specific Death | 17.56 (0.01) | 17.27 (0.01) | 19.23 (0.01) |  |
| <sup>1</sup> chi-squared test with Rao & Scott's second-order correction |  |  |  |  |
| Note: CVD- Cardiovascular |  |  |  |  |

